# Continuity of Care by Primary Care Provider in Young Children with Chronic Conditions

**DOI:** 10.1101/2021.02.04.21251018

**Authors:** Yair Bannett, Rebecca M. Gardner, Lynne C. Huffman, Heidi M. Feldman, Lee M. Sanders

**Author notes:** **Address correspondence to:** Yair Bannett, Developmental-Behavioral Pediatrics, Stanford University School of Medicine, 1265 Welch Road, X109, Stanford, CA, 94305. **Financial Disclosure:** The authors have no financial relationships relevant to this article to disclose.

## Abstract

**Objectives:** (1) To assess continuity of care by primary-care provider (CoC), an established quality indicator, in children with asthma, autism spectrum disorder (ASD), and no chronic conditions, and (2) to determine patient factors that influenced CoC.

**Methods:** Retrospective cohort study of electronic health records from all office visits of children under 9 years, seen ≥4 times between 2015 and 2019 in 10 practices of a community-based primary healthcare network in California. Three cohorts were constructed: (1)Asthma: ≥2 visits with asthma visit diagnoses; (2)ASD: same method; (3)Controls: no chronic conditions. CoC, using the Usual Provider of Care measure (range >0-1), was calculated for (1)total visits and (2)well-care visits only. Fractional regression models examined CoC adjusting for patient age, medical insurance, practice affiliation, and number of visits.

**Results:** Of 30,678 eligible children, 1875 (6.1%) were classified as Asthma, 294 (1.0%) as ASD, and 15,465 (50.4%) as Controls. Asthma and ASD had lower total CoC than Controls (Mean=0.58, SD 0.21, M=0.57, SD 0.20, M=0.66, SD 0.21). Differences among well-care CoC were smaller (Asthma M=0.80, ASD M=0.78, Controls M=0.82). In regression models, lower total CoC was found for Asthma (aOR 0.90, 95% CI 0.85-0.94). Lower total and well-care CoC were associated with public insurance (aOR 0.77, CI 0.74-0.81; aOR 0.64, CI 0.59-0.69).

**Conclusion:** Children with asthma in this primary-care network had lower CoC compared to children without chronic conditions. Public insurance was the most prominent patient factor associated with low CoC. Quality initiatives should address disparities in CoC for children with chronic conditions.

**Table of Contents Summary:** Continuity of care by primary care provider is an established quality indicator. We compared continuity in young children with asthma, autism, and no chronic conditions.

**What’s Known on This Subject:** Continuity of care has emerged as an important component of care in the patient-centered medical home, especially for children with chronic medical conditions. However, it has been minimally studied across chronic conditions, especially in neurodevelopmental disorders.

**What This Study Adds:** Children with asthma, but not those with autism spectrum disorder, had lower continuity of care compared to children without chronic conditions. Public insurance was associated with lower care continuity for children with and without chronic conditions, highlighting important sociodemographic disparities.

**Contributors’ Statement Page:** Dr. Bannett conceptualized and designed the study, defined and coordinated data extraction, carried out the data analyses, drafted the manuscript, and reviewed and revised the manuscript.

Ms. Gardner participated in study design, extensively reformatted the data for analysis, performed statistical data analysis, and critically reviewed and revised the manuscript.

Dr. Feldman participated in study design, supervised data analysis and critically reviewed and revised the manuscript.

Drs. Huffman and Sanders supervised the conceptualization and design of the study, supervised data analysis, and critically reviewed and revised the manuscript.

All authors approved the final manuscript as submitted and are responsible for all aspects of the work.

## Introduction

Continuity of care by primary care provider (CoC) is widely considered a core component of the patient-centered medical home. CoC is an established quality indicator that has been linked – especially in children with chronic conditions – to improved care coordination, healthcare utilization, and receipt of preventive care.^1-6^ However, in current primary care practice, children may not be seen by the same primary care provider (PCP) in their first years of life, potentially contributing to over- and under treatment, compromising patient safety, and ultimately adversely affecting health outcomes. To date, the extent of CoC has not been compared across different pediatric chronic conditions and has rarely been evaluated in children with neurodevelopmental disorders.

Autism spectrum disorder and asthma are common chronic conditions of childhood, requiring continuous care management by a skilled primary care provider. Asthma affects 8-9% of US children,^7^ and ambulatory asthma care is mainly provided in primary care settings.^8^ High continuity of ambulatory asthma care in children has been shown to be associated with decreased asthma-specific emergency department utilization and number of hospitalizations.^1,9-11^ Autism spectrum disorder (ASD) is a chronic neurodevelopmental condition estimated to affect approximately 2% of US children.^12,13^ The American Academy of Pediatrics (AAP) published guidelines in 2001 and 2006 stating that the primary care medical home is the ideal setting for developmental screening and identification of early signs of ASD.^14-16^ CoC might be especially important in young children with ASD, given its potential influence on timely diagnosis and treatment of ASD, which have been shown to improve patient outcomes.^17,18^ However, CoC has not been studied in young children with ASD.

Comparing CoC in children with physical and neurodevelopmental chronic conditions, and identifying factors associated with low CoC, may inform targeted quality improvement interventions aimed at improving high-quality care of children with chronic conditions. In this study, we examined electronic health record (EHR) data from a large community-based pediatrics primary care network (1) to compare CoC in young children with asthma and ASD to children with no chronic condition, and (2) to determine patient factors that influence continuity of care. We hypothesized that CoC would be low in children with asthma and ASD compared to those without chronic conditions and that differences in CoC between children with asthma and ASD would be accounted for by number of primary care visits.

## Methods

### Setting

Packard Children’s Health Alliance (PCHA) is a community-based pediatric healthcare network in the San Francisco Bay Area, affiliated with Stanford Children’s Health and Lucile Packard Children’s Hospital. PCHA has 24 pediatrics primary care offices, grouped into 10 practices.

### Data Sources and Population

We conducted a retrospective review of EHRs for a cohort of all pediatric patients under the age of 9 years seen by PCHA PCPs from October 1, 2015 (date coinciding with adoption of ICD-10 codes) to December 31, 2019. We extracted de-identified structured data from all office encounters. To create a robust and meaningful measure of CoC, we included only children who had at least 4 visits during the examined period and who were seen over a minimum of one year. Figure 1 shows the study cohort flowchart.

**Figure 1.**
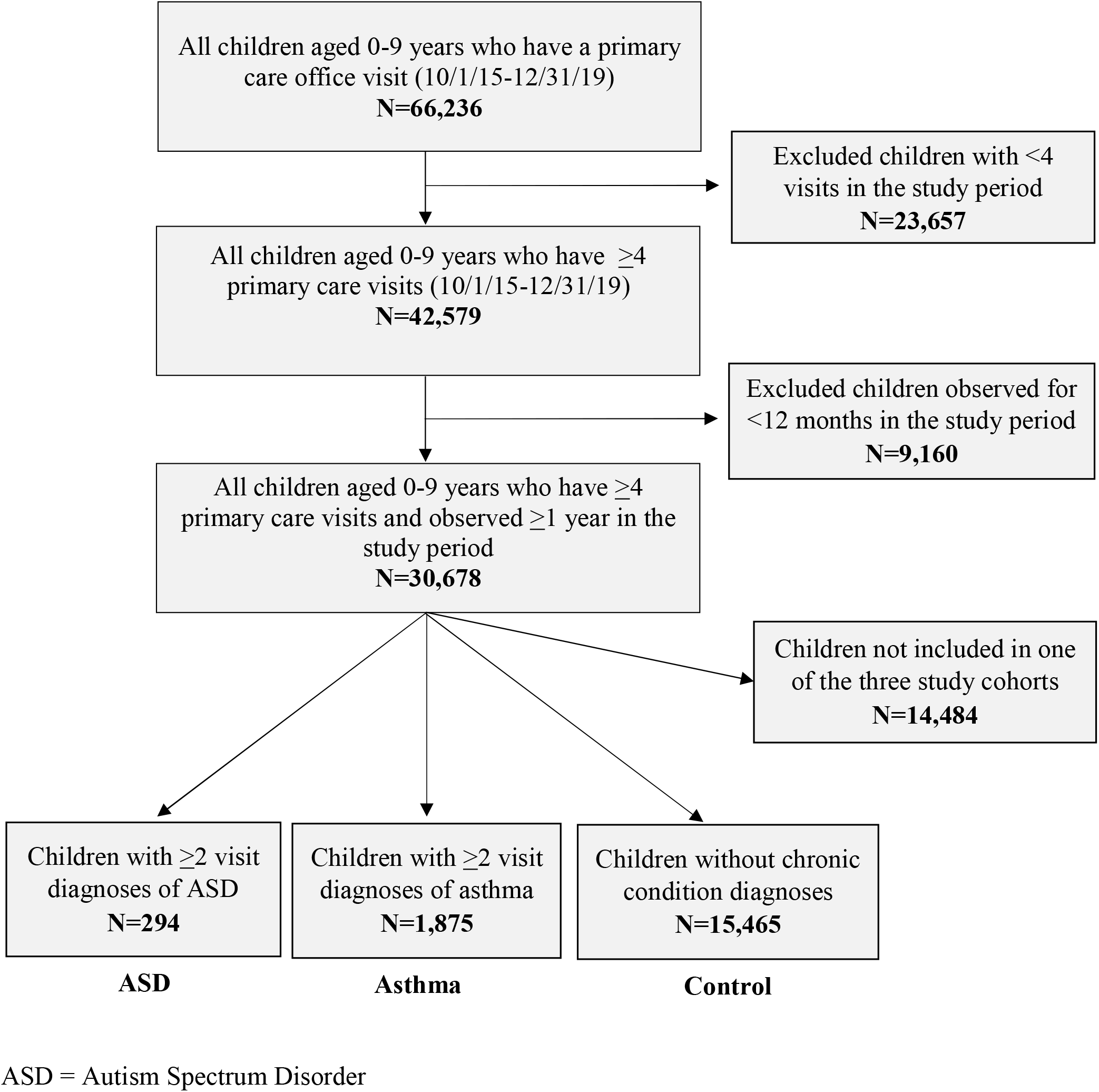
Study Flow Diagram.

This study was determined by the Stanford University School of Medicine institutional review board to not represent human subject research.

### Measures

### Study outcomes: Continuity of Care by Primary Care Provider

We calculated continuity of care (CoC) using the Usual Provider of Care (UPC) measure (range: >0 - 1), an easily interpretable measure that is commonly used in EHR-based studies that examine CoC.^19^ This measure is calculated as a ratio: for each patient, the number of primary care visits conducted by the most commonly seen provider is divided by the total number of primary care visits. As done in previous studies, we calculated two CoC outcomes per patient: (1) total, based on all visits and (2) well-care, based on well-child visits only.^5^ We defined well-care visits as visits with a code or descriptor representing a well-care visit in at least one of the following structured EHR fields: Visit diagnosis code; Current Precedural Terminology (CPT) code; Visit type descriptor (see Supplementary 1). We also used the Visit type structured field to define same day sick visits as visits that had the descriptor “same day sick”.

### Primary independent variable: Study Cohorts

We constructed the following 3 cohorts: (1) Asthma – included patients who had at least two visits in the study period with a visit diagnosis code of asthma (and no ASD diagnoses); (2) ASD - included patients who had at least two visits in the study period with a visit diagnosis code of Autism Spectrum Disorder; (3) Controls – patients who did not have any chronic condition diagnosed during the study period, based on a list of pediatric chronic conditions.^20^ (see Supplementary 1).

### Secondary Independent Variables

Health care utilization factors: acknowledging that the vast majority of patients in this network (97%) received their care during the study period solely in one primary care practice, we compared CoC across the 10 practices and included the primary care practice affiliation as an independent variable in regression models. The number of visits per patient during the study period, which has a direct impact on the ability to maintain CoC, was another important independent variable we included in regression models.

We used patient structured data in the EHR to describe the following patient characteristics: patient age (mean age in years during study period), sex, race (White/Asian/African American/Other), ethnicity (Hispanic/Non-Hispanic), and medical insurance at first study period visit (private/public/military).

### Statistical Analysis

We used descriptive statistics and standardized mean differences (SMD) to summarize and compare patient characteristics across the three cohorts.^21^ SMD values of 0.2, 0.5, and 0.8 correspond to small, moderate, and large differences, respectively. To examine associations among cohort affiliation, patient characteristics, and study outcomes, we implemented fractional logistic regression models and calculated 95% confidence intervals for the adjusted odds ratios (aOR). Patient race and ethnicity data were not included in regression models due to high percent of missing data (32% for race and 35% for ethnicity). Imputation was not attempted, since race/ethnicity data were highly correlated with insurance type (see Supplementary 2); therefore, the addition of an imputed race/ethnicity variable would likely not yield additional information beyond what was provided by insurance type. Patient sex was also not included in regression models because of the high prevalence of ASD in males and lack of an underlying hypothesis to link patient sex with continuity of care in young children.

Model 1 compared CoC (total and well-care only) across the three patient cohorts while adjusting for patient age, medical insurance, practice affiliation, and number of visits. To mitigate the observed difference in age distribution across the cohorts, likely stemming from the fact that both ASD and asthma are usually not diagnosed in the first 2 years of life, we performed a sensitivity analysis in which we included only patients who reached a minimum age of 2 years during the study period (Model 2). To further explore the influence of insurance type on total continuity of care, a post-hoc analysis was completed, in which the regression model was implemented after patients were stratified based on insurance type (Model 3). Since CoC was not normally distributed across this finite sample, we recognized that asymptotic normality might not hold.^22,23^ Therefore, we used bootstrapping to calculate the standard errors of every model for valid inference.^24^ All analyses were conducted using R, version 3.6.2.^25^

## Results

Of 30,678 eligible children aged 0 to 8 years, 1,875 (6.1%) were classified with Asthma, 294 (1.0%) with ASD, and 15,465 (50.4%) as Controls. Table 1 describes patient and clinical care characteristics of the three cohorts. Patients in the Control cohort were younger than the Asthma and ASD cohorts (standardized mean difference (SMD)=0.44). The ASD cohort was predominantly male (78.2%); both Asthma and ASD had a higher percent of publicly insured patients (25.3% and 23.8%) compared to Controls (14.9%). The number of patients that switched between private and public insurance during the study period did not differ across the three cohorts (Asthma=77 (4.1%), ASD=11 (3.7%), Controls=371 (2.4%), SMD=0.06). Total visits in the study period for Asthma averaged 17.4 compared to 14.0 for ASD and 11.4 for Controls (SMD=0.41). Number of same day sick visits did not differ across the cohorts (Asthma=11,336 (45.9%), ASD=1,301 (44.9%), Controls=39,989 (43.0%), SMD=0.04).

**Table 1.** Patient and clinical care characteristics of study cohorts aged 0-8 years

|  | Control | Asthma | ASD | Standardized Mean Difference |
| --- | --- | --- | --- | --- |
| **n** | **15,465** | **1,875** | **294** |  |
| Age<sup>a</sup> | 3.6 (2.3) | 5.0 (1.9) | 4.9 (1.8) | 0.44 |
| Male (%) | 7,314 (47.3) | 1,137 (60.6) | 230 (78.2) | 0.45 |
| Insurance<sup>b</sup> (%) |  |  |  | 0.23 |
| Private | 12,395 (81.1) | 1,349 (72.3) | 202 (68.9) |  |
| Public | 2,270 (14.9) | 463 (24.8) | 74 (25.3) |  |
| Military | 617 (4.0) | 54 (2.9) | 17 (5.8) |  |
| Insurance switch<sup>c</sup> | 371 (2.4) | 77 (4.1) | 11 (3.7) | 0.06 |
| Race (%) |  |  |  | 0.24 |
| White | 4,411 (28.5) | 479 (25.5) | 67 (22.8) |  |
| Asian | 2,793 (18.1) | 347 (18.5) | 67 (22.8) |  |
| Black | 387 (2.5) | 122 (6.5) | 10 (3.4) |  |
| Other<sup>d</sup> | 2,292 (14.8) | 407 (21.7) | 59 (20.1) |  |
| Unknown | 5,582 (36.1) | 520 (27.7) | 91 (31.0) |  |
| Ethnicity (%) |  |  |  | 0.17 |
| Hispanic | 1,331 (8.6) | 273 (14.6) | 41 (13.9) |  |
| Not hispanic | 8,437 (54.6) | 1,085 (57.9) | 160 (54.4) |  |
| Unknown | 5,697 (36.8) | 517 (27.6) | 93 (31.6) |  |
| Total Visits<sup>e</sup> | 11.4 (7.4) | 17.4 (11.7) | 14.0 (9.6) | 0.41 |
| Well-Care Visits<sup>f</sup> | 5.4 (3.5) | 4.2 (2.9) | 4.1 (2.9) | 0.28 |
| Same Day Sick Visits (%)<sup>g</sup> | 39,989 (43.0) | 11,336 (45.9) | 1,301 (44.9) | 0.04 |
| Total Continuity of Care | 0.66 (0.21) | 0.57 (0.20) | 0.58 (0.21) | 0.26 |
| Well-Care Continuity of Care | 0.82 (0.22) | 0.80 (0.23) | 0.78 (0.23) | 0.11 |

<sup>a</sup>Age in years at midpoint in study. Mean and standard deviation is displayed for continuous variables, including age, total visits, well-care visits, same day sick visits, total continuity of care, and well-care continuity of care.
<sup>b</sup>Insurance type in the first visit during the study period.
<sup>c</sup>Patient switched between private and public insurance at least once in the study period
<sup>d</sup>Includes Other, Native American and Pacific Islander.
<sup>e</sup>Total number of all office visits during the study period.
<sup>f</sup>Number of Well-Care visits during the study period.
<sup>g</sup>Number of Same Day Sick Visits (% of all non-Well-Care Visits).
ASD = Autism Spectrum Disorder

Total CoC as measured by the UPC score was lower in children with Asthma and ASD (Mean (M)=0.57, Standard Deviation (SD) 0.20; M=0.58, SD 0.21) compared to Controls (M=0.66, SD 0.21); SMD=0.26. Differences among well-care CoC were less pronounced (Asthma M=0.80, ASD M =0.78, Control M=0.82; SMD=0.11).

To examine differences in total CoC across the 10 primary care practices, we stratified patient UPC scores by practice. Figure 2a illustrates the high variation in total CoC across the practices, ranging from an average UPC of 0.5 in three practices to 0.78 in one practice. The number of PCPs in each practice ranged from 6 to 37. The number of PCPs per practice was positively correlated with the practice UPC score, but the correlation did not reach statistical significance (R=0.59, 95% CI −0.89, −0.06, p=0.071). In Figure 2b, we further stratified total CoC by patient cohort (Asthma, ASD, Controls), demonstrating a persistent pattern of total CoC across patient cohorts in most practices (Asthma<ASD<Controls). Practices that did not follow this pattern had notably small cohort groups for ASD or Asthma patients (n<20). In Figure 2c, we stratified patients by insurance type (public vs. private insurance), given the higher percent of publicly insured patients in Asthma and ASD compared to Controls. This figure demonstrates that within each of the 5 practices that had a substantial proportion of publicly insured patients, these patients had consistently lower total CoC compared to patients with private insurance; In two of the 5 practices, the UPC score for publicly insured patients was below 0.5.

**Figure 2a.**
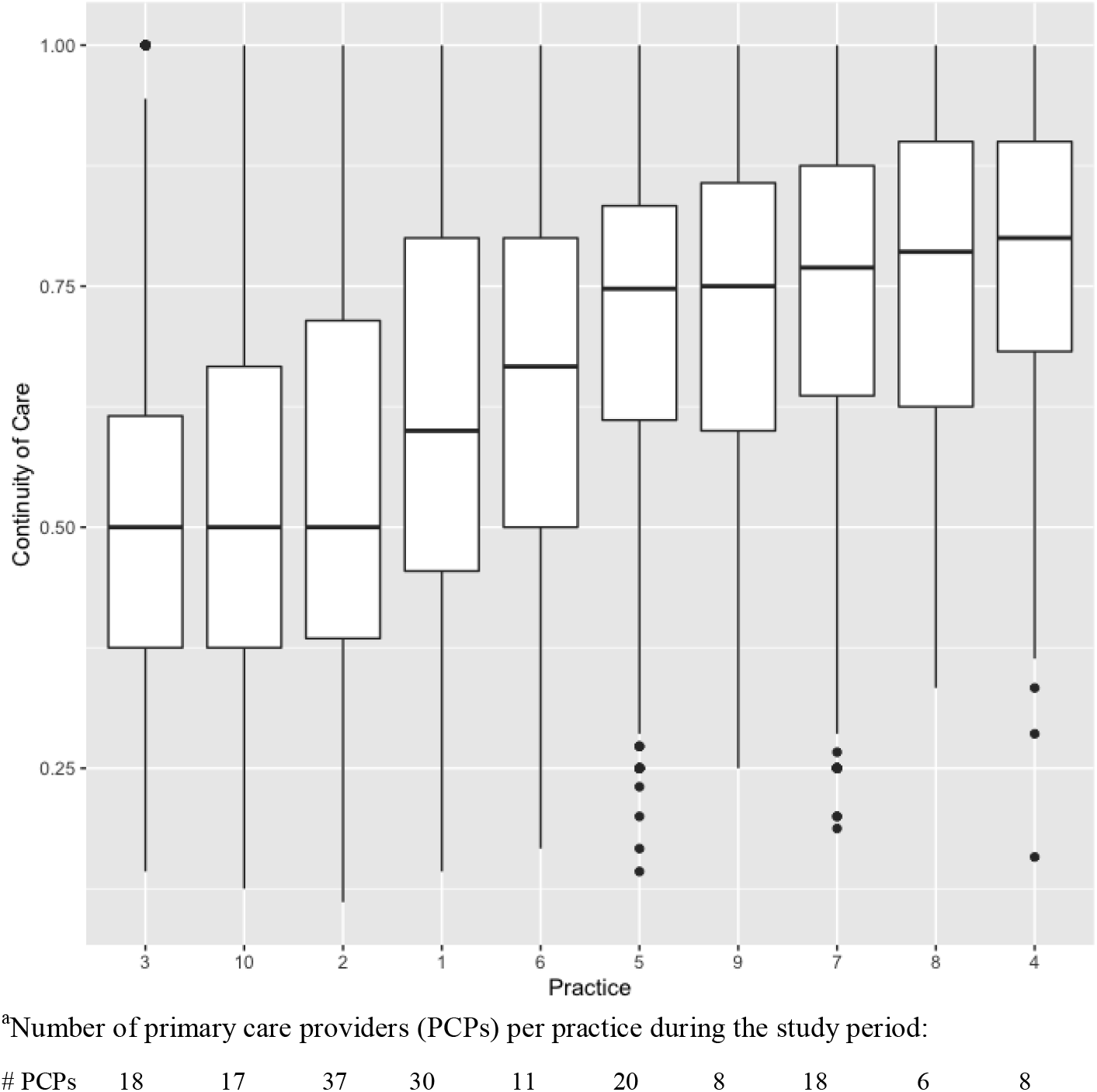
Continuity of Care by Practice.^a^

**Figure 2b.**
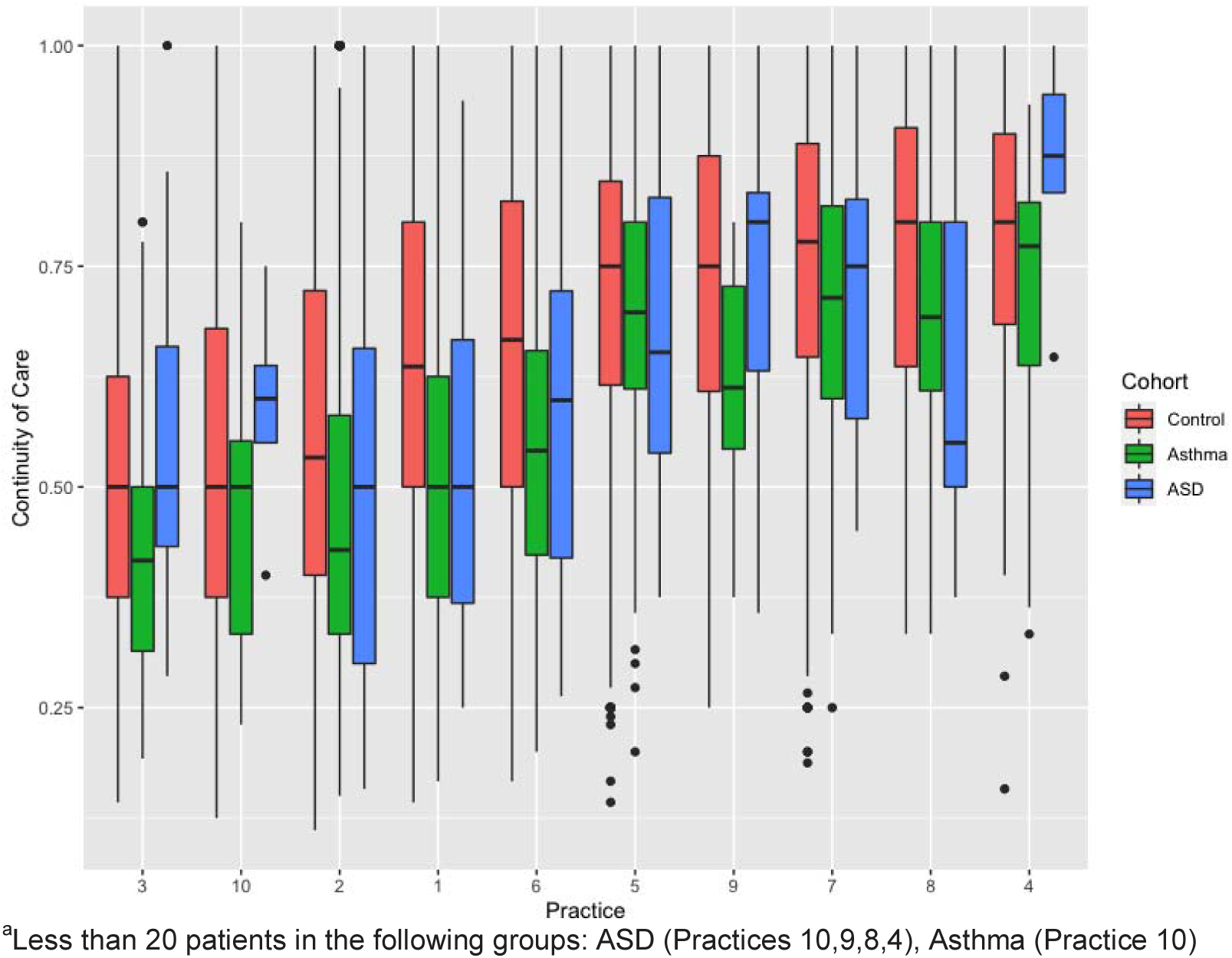
Continuity of Care by Practice, stratified by patient cohort.^a^

**Figure 2c.**
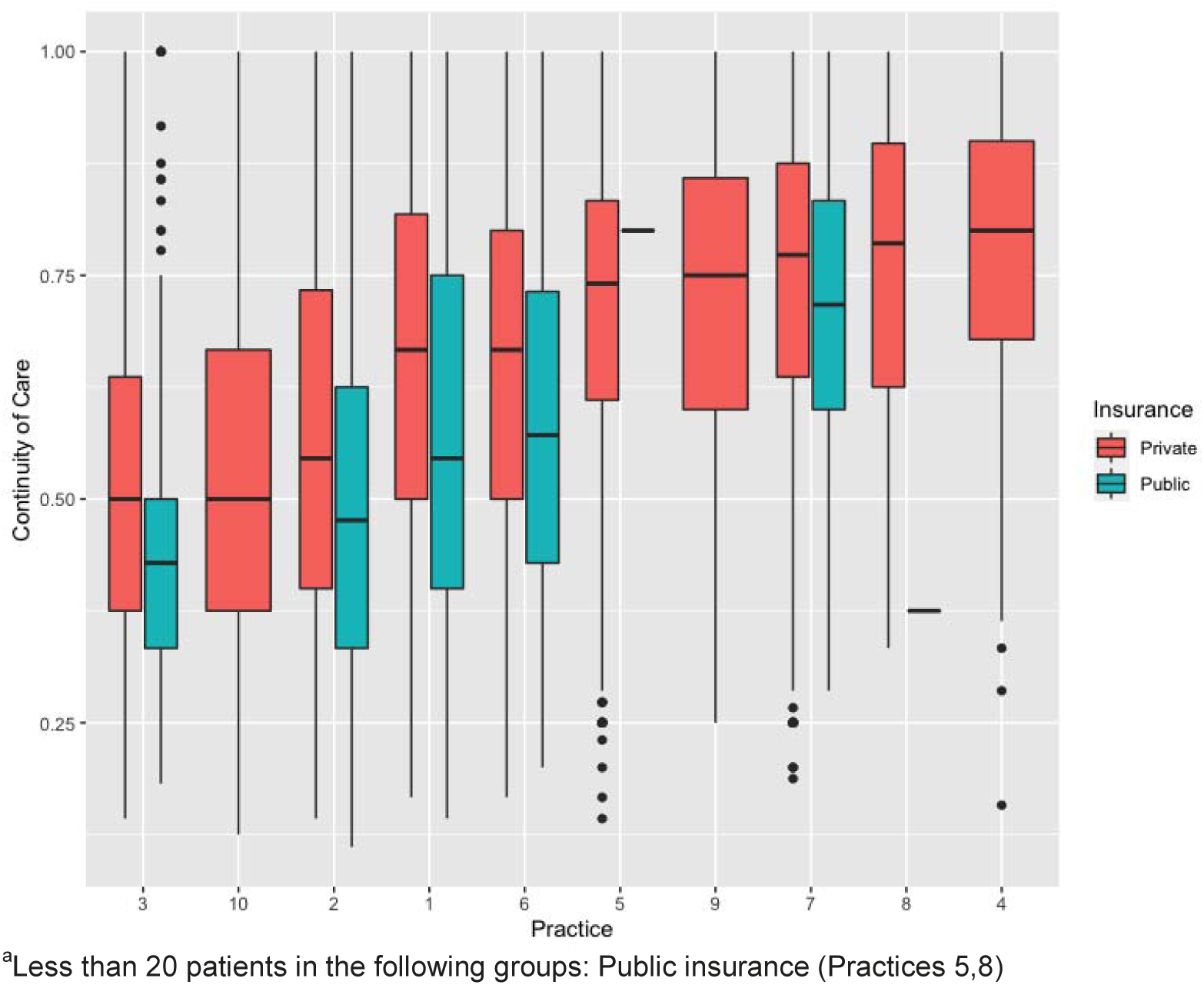
Continuity of Care by Practice, stratified by insurance.

### Regression models

Table 2 shows the results of regression models for total and well-care CoC. After accounting for patient age, practice affiliation, and number of visits in the study period, lower total CoC was found for Asthma (aOR 0.90, 95% CI 0.85-0.94) compared to Controls and was associated with having public or military insurance (aOR 0.77, CI 0.74-0.81; aOR 0.92, CI 0.85-0.99) compared to private insurance. The difference in total CoC between ASD and Controls did not reach statistical significance. Well-care CoC did not substantively differ across the cohort groups.

**Table 2.** Resuts of fractional regression models comparing continuity of care across patient cohorts and insurance type, adjusting for patient age, number of visits, and practice affiliation

| **Total Continuity of Care** | aOR | Lower CI | Upper CI | P value |
| --- | --- | --- | --- | --- |
| Study Cohort (ref=Control) |  |  |  |  |
| ASD | 0.94 | 0.84 | 1.04 | 0.254 |
| Asthma | 0.90 | 0.86 | 0.94 | <0.001 |
| Insurance (ref=Private) |  |  |  |  |
| Public | 0.77 | 0.74 | 0.81 | <0.001 |
| Military | 0.92 | 0.85 | 0.99 | 0.026 |
| **Well-Care Continuity of Care** | aOR | Lower CI | Upper CI | P value |
| Study Cohort (ref=Control) |  |  |  |  |
| ASD | 0.96 | 0.81 | 1.12 | 0.650 |
| Asthma | 0.97 | 0.90 | 1.04 | 0.472 |
| Insurance (ref=Private) |  |  |  |  |
| Public | 0.64 | 0.59 | 0.69 | <0.001 |
| Military | 0.94 | 0.81 | 1.07 | 0.345 |

aOR = Adjusted Odds Ratio, CI = Confidence Interval (95%).

However, low well-care CoC was associated with public insurance (aOR 0.64, CI 0.59-0.69). A sensitivity analysis, adding a requirement that patients reach the age of 2 years to be included in the study, did not change any of the results substantively (Supplementary 3).

### Post-hoc analyses

To further explore the strong association between patient public insurance and low total CoC, we implemented post-hoc regression models stratified by insurance type. Table 3 shows the results of three regression models, one for each insurance type (i.e., private, public, military).

**Table 3.** Post-hoc fractional regression models stratified by insurance type, comparing total continuity of care across patient cohorts, adjusting for patient age, number of visits, and practice affiliation.

| **Private Insurance** | aOR | Lower CI | Upper CI | P value |
| --- | --- | --- | --- | --- |
| Study Cohort (ref=Control) |  |  |  |  |
| ASD | 0.91 | 0.79 | 1.03 | 0.11 |
| Asthma | 0.89 | 0.84 | 0.94 | <0.001 |
| **Public Insurance** | aOR | Lower CI | Upper CI | P value |
| Study Cohort (ref=Control) |  |  |  |  |
| ASD | 1 | 0.83 | 1.18 | 0.959 |
| Asthma | 0.9 | 0.82 | 0.98 | 0.011 |
| **Military Insurance** | aOR | Lower CI | Upper CI | P value |
| Study Cohort (ref=Control) |  |  |  |  |
| ASD | 1.33 | 0.98 | 1.68 | 0.11 |
| Asthma | 1.07 | 0.85 | 1.3 | 0.545 |

aOR = Adjusted Odds Ratio, CI = Confidence Interval (95%).

Lower total CoC for Asthma as compared to Controls persisted within the privately and publicly insured cohorts (aOR 0.89, CI 0.84-0.94; OR 0.90, CI 0.82-0.98), but did not persist in military insured patients (aOR 1.07, CI 0.85-1.30). Military insured patients also had less variation in total CoC by practice (Table 3).

## Discussion

This EHR-based examination of continuity of care (CoC) in a community-based pediatrics primary healthcare network, revealed that children diagnosed with asthma and with autism-spectrum disorder (ASD) had lower total CoC, compared to children without chronic conditions. After accounting for patient age, practice affiliation and number of visits, lower total CoC was found in the Asthma cohort, but not the ASD cohort. Lower total and lower well-care CoC each were associated with having public insurance. In post-hoc analyses, after stratifying patients by insurance type, the Asthma cohort persistently had lower total CoC.

Absolute differences in total CoC across the cohorts showed that children with ASD and asthma saw their usual PCP on average 6/10 times, while children without chronic conditions saw their usual PCP on average 7/10 times. This finding, though modest in magnitude, aligns with previous literature that found low total CoC in children with chronic and complex medical conditions.^1,3^ The high well-care CoC across the primary-care network (mean UPC score ∼0.8), as well as the minimal differences across the three study cohorts, could reflect a delibarate effort by health organizations, PCPs, and patients to maintain CoC for childhood preventive visits. It is important to note, however, that in prior studies, total CoC, but not well-care CoC, was associated with improved healthcare utilization and receipt of preventive care.^2,5^ On the other hand, in children with chronic conditions, well-care attendance has been associated with lower risk of ambulatory care-sensitive hospitalizations, and in children with asthma was linked to reductions in asthma exacerbations.^3,26^ Future studies should assess whether in children with ASD, attendance at well-child care visits and/or well-care CoC are associated with favorable patient outcomes (e.g., timely diagnosis and referral to treatment).

In fractional regression models, patients with Asthma, but not those with ASD, had statistically significant lower total CoC compared to Controls. Contrary to our hypothesis, differences in CoC between children with Asthma and those with ASD persisted after accounting for number of primary care visits. Realizing CoC is not the same across two chronic conditions, we tried to understand if there are the unique condition-specific factors that drive differences in CoC for children with these two distinct chronic conditions. For patients with asthma, we considered the urgent nature of visits as well as PCP and family perceptions of the condition. To examine the potential influence of the urgent nature of some primary care visits of children with asthma, we compared the rates of same day sick visits across study cohorts, but did not find a substantial difference. Prior literature found that many parents of children with asthma consider asthma to be an “episodic” condition, which has been recognized as one explanation for lack of patient adherence to asthma controller medication.^27,28^ Further research is needed to assess whether this perception also has an effect on the family’s effort to maintain continuous care with the same clinician. For patients with ASD, who represented the smallest cohort, CoC varied across practices, as illustrated in Figure 2b. In four of the ten practices patients with ASD had equal or higher CoC scores compared to Controls. Different from asthma, families of children with ASD and their PCPs may have a stronger perceived need to maintain continuity of care for these children. Such condition-specific differences are important to further explore in an effort to target specific patient populations that require interventions to enhance CoC.

Our study confirms that reduced CoC for children with chronic conditions does not extend to a neurodevelopmental disorder, ASD, after accounting for differences in patient characteristics and health care utilization across the cohorts. Such differences should be explored in other chronic conditions. It will also be critical to identify condition-specific factors the that may impede CoC as well as the moderating influence of social factors – including parent language and ethnicity and neighborhood. Such factors can then become targets for interventions aimed at enhancing high-quality care delivery to all children with chronic physical and mental health conditions.

The strong association we found through regression models between public insurance and low continuity of care, across all patient cohorts and for both total and well-care CoC, raises concerns for sociodemographic disparities in quality of care. Previous studies in other primary health care systems also found a similar pattern of low CoC in publicly insured patients.^1,5^ Systems-level barriers present one possible explanation for this phenomenon. The mandatory requirement to renew public insurance annualy and the limited number of providers that accept publicly insured patients may prevent children with public insurance from receiving optimal care. Empirical evidence to support this explanation can be found in a study that found that PCP availability was the most significant factor associated with CoC in primary care settings, much more than family factors, such as perceived importance of continuity.^29^ In an attempt to further explore the association between public insurance and low continuity of care and its influence on continuity of care across patient cohorts, we implemented post-hoc regression models. By stratifying by insurance type, we learned that differences in CoC across patient cohorts persisted within publicly and privately insured patients, suggesting there are condition-specific factors that influence CoC, resulting in lower CoC in children with asthma than in children with a neurodevelopmental chronic condition, ASD. The lack of difference in CoC across patient cohorts and the reduced variability in CoC across practices in patients with military insurance could represent a more consistent rate of access to primary care providers for all patients served in this system.

Thus, we found that use of public insurance was a consistent contributor to reduced CoC across the two chronic conditions. It is important to recognize that disparities in CoC represent disparities in quality of care for children with chronic conditions and that these disparities may compromise health outcomes in an already burdened group. To successfully mitigate these disparities, studies that differentiate between patient and family factors (e.g., language proficiency), clinician and practice factors (e.g., availability of interpreting services), and health system (e.g., insurance type) factors that drive CoC may identify modifiable factors in CoC.

To our knowledge, this study is the first to compare CoC across two disparate but common pediatric chronic conditions. The unexpected finding Several limitations should be acknowledged. The cross-sectional study design did not allow assessment of causality and the potential influence of CoC on patient outcomes. As such, we were limited in our ability to assess the clinical significance of the differences we found in CoC across patient cohorts. PCHA is a population-based practice network, but it represents one healthcare system, which limits generalizability. Finally, use of structured EHR data and the significant proportion of missing data on patient race/ethnicity limited the evaluation of important patient factors that may contribute to disparities in CoC.

## Conclusion

This EHR-based study of a large community-based pediatrics primary care network demonstrated disparities in continuity of care (CoC), an important quality of care indicator for children with chronic conditions, such as asthma and autism-spectrum disorder. The strong association we found between public insurance and low CoC highlights the need to explore system-level and condition-specific drivers that should be targeted in an effort to mitigate disparities in quality of health care provided to children with chronic conditions.

## Data Availability

Datasets used for this study must be completely de-identified according to federal regulations prior to release for sharing. Given the potentially sensitive nature of these data, we will make the datasets available only under a data-sharing plan. Requests to use the information and data from this study will be considered on a case-by-case basis, following written request to the corresponding author.

## Abbreviations

AAP: American Academy of Pediatrics

ADHD: Attention-Deficit/Hyperactivity Disorder

ASD: Autism Spectrum Disorder

CoC: Continuity of Care

EHR: Electronic Health Record

PCHA: Packard Children’s Health Alliance

PCP: Primary Care Provider

## Acknowledgments

We thank the members of the Stanford Quantitative Sciences Unit for their review and feedback on the manuscript. We thank Packard Children’s Health Alliance and Stanford Research Information Technology for their support and assistance in data acquisition and extraction.

Completion of this study is a fulfillment of one of the requirements for Dr. Bannett’s master’s degree in Health Research and Policy at Stanford University.

## References

1. Christakis DA, Mell L, Koepsell TD, Zimmerman FJ, Connell FA. Association of lower continuity of care with greater risk of emergency department use and hospitalization in children. Pediatrics. 2001;107(3):524–529.

2. Flores AI, Bilker WB, Alessandrini EA. Effects of continuity of care in infancy on receipt of lead, anemia, and tuberculosis screening. Pediatrics. 2008;121(3):e399–406.

3. Tom JO, Tseng CW, Davis J, Solomon C, Zhou C, Mangione-Smith R. Missed well-child care visits, low continuity of care, and risk of ambulatory care-sensitive hospitalizations in young children. Archives of pediatrics & adolescent medicine. 2010;164(11):1052–1058.

4. Chen AY, Schrager SM, Mangione-Smith R. Quality measures for primary care of complex pediatric patients. Pediatrics. 2012;129(3):433–445.

5. Enlow E, Passarella M, Lorch SA. Continuity of Care in Infancy and Early Childhood Health Outcomes. Pediatrics. 2017;140(1).

6. Arthur KC, Mangione-Smith R, Burkhart Q, et al. Quality of Care for Children With Medical Complexity: An Analysis of Continuity of Care as a Potential Quality Indicator. Academic pediatrics. 2018;18(6):669–676.

7. Akinbami LJ, Simon AE, Rossen LM. Changing Trends in Asthma Prevalence Among Children. Pediatrics. 2016;137(1):1–7.

8. Akinbami LJ, Santo L, Williams S, Rechtsteiner EA, Strashny A. Characteristics of Asthma Visits to Physician Offices in the United States: 2012-2015 National Ambulatory Medical Care Survey. National health statistics reports. 2019(128):1–20.

9. Cree M, Bell NR, Johnson D, Carriere KC. Increased continuity of care associated with decreased hospital care and emergency department visits for patients with asthma. Dis Manag. 2006;9(1):63–71.

10. Dombkowski KJ, Harrison SR, Cohn LM, Lewis TC, Clark SJ. Continuity of prescribers of short-acting beta agonists among children with asthma. J Pediatr. 2009;155(6):788–794.

11. Huang ST, Wu SC, Hung YN, Lin IP. Effects of continuity of care on emergency department utilization in children with asthma. Am J Manag Care. 2016;22(1):e31–37.

12. Baio J, Wiggins L, Christensen DL, et al. Prevalence of Autism Spectrum Disorder Among Children Aged 8 Years - Autism and Developmental Disabilities Monitoring Network, 11 Sites, United States, 2014. Morbidity and mortality weekly report Surveillance summaries (Washington, DC : 2002). 2018;67(6):1–23.

13. Kogan MD, Vladutiu CJ, Schieve LA, et al. The Prevalence of Parent-Reported Autism Spectrum Disorder Among US Children. Pediatrics. 2018;142(6).

14. American Academy of Pediatrics: The pediatrician’s role in the diagnosis and management of autistic spectrum disorder in children. Pediatrics. 2001;107(5):1221–1226.

15. Identifying infants and young children with developmental disorders in the medical home: an algorithm for developmental surveillance and screening. Pediatrics. 2006;118(1):405–420.

16. Johnson CP, Myers SM. Identification and evaluation of children with autism spectrum disorders. Pediatrics. 2007;120(5):1183–1215.

17. Zwaigenbaum L, Bauman ML, Stone WL, et al. Early Identification of Autism Spectrum Disorder: Recommendations for Practice and Research. Pediatrics. 2015;136 Suppl 1:S10–40.

18. Vivanti G, Dissanayake C. Outcome for Children Receiving the Early Start Denver Model Before and After 48 Months. Journal of autism and developmental disorders. 2016;46(7):2441–2449.

19. Breslau N, Reeb KG. Continuity of care in a university-based practice. J Med Educ. 1975;50(10):965–969.

20. Mokkink LB, van der Lee JH, Grootenhuis MA, Offringa M, Heymans HS. Defining chronic diseases and health conditions in childhood (0-18 years of age): national consensus in the Netherlands. Eur J Pediatr. 2008;167(12):1441–1447.

21. Austin PC. Using the Standardized Difference to Compare the Prevalence of a Binary Variable Between Two Groups in Observational Research. Communications in Statistics - Simulation and Computation. 2009;38(6):1228–1234.

22. Rascati KL, Smith MJ, Neilands T. Dealing with skewed data: an example using asthma-related costs of medicaid clients. Clin Ther. 2001;23(3):481–498.

23. Visalakshi J, Jeyaseelan L. Confidence Interval for skewed distribution in outcome of change or difference between methods. Clinical Epidemiology and Global Health. 2014;2(3):117–120.

24. Efron B, Tibshirani RJ. An introduction to the bootstrap. CRC press; 1994.

25. R: A language and environment for statistical computing. [computer program]. R Foundation for Statistical Computing, Vienna, Austria. 2018.

26. Lang JE, Tang M, Zhao C, Hurst J, Wu A, Goldstein BA. Well-Child Care Attendance and Risk of Asthma Exacerbations. Pediatrics. 2020.

27. Peláez S, Bacon SL, Lacoste G, Lavoie KL. How can adherence to asthma medication be enhanced? Triangulation of key asthma stakeholders’ perspectives. J Asthma. 2016;53(10):1076–1084.

28. Mowrer JL, Tapp H, Ludden T, et al. Patients’ and providers’ perceptions of asthma and asthma care: a qualitative study. J Asthma. 2015;52(9):949–956.

29. Christakis DA, Kazak AE, Wright JA, Zimmerman FJ, Bassett AL, Connell FA. What factors are associated with achieving high continuity of care? Family medicine. 2004;36(1):55–60.

