## Supplemental tables for "Continuity of Care by Primary Care Provider in Young Children with Chronic Conditions"

**Supplement**

**Supplementary 1.** Codes and descriptors used for cohort definitions and Well-Care Visit

**Supplementary 2**. Association between race/ethnicity and insurance type

**Supplementary 3.** Sensitivity analysis with age restriction (minimum required age of 2 years)

**Supplementary 1.** Codes and descriptors used for cohort definitions and Well-Care Visit

|  | EHR Field | | |
| --- | --- | --- | --- |
|  | **ICD-10 Diagnosis codes** | **Current Procedural Terminology (CPT) codes** | **Visit Type** |
| ASD | F84.X F94.8 F94.9 F80.82 | NA | NA |
| Asthma | J45.X | NA | NA |
| Well-Care Visit | Z00.129 Z00.121 Z00.00 | 99381 99382 99383 99384 99391 99392 99393 99394 99395 99401 99402 | Well-Care Visit (and other similar descriptors) |

ASD = Autism Spectrum Disorder.

**Supplementary 2**. Association between race/ethnicity and insurance type

|  | Private | Public | Military | P value |
| --- | --- | --- | --- | --- |
| n | 13,946 | 2,807 | 688 |  |
| Race (%) |  |  |  | <0.001 |
| Asian | 4,366 (31.3) | 448 (16.0) | 103 (15.0) |  |
| Black | 191 (1.4) | 313 (11.2) | 6 (0.9) |  |
| Other | 2,896 (20.8) | 269 (9.6) | 13 (1.9) |  |
| Unknown | 1,708 (12.2) | 974 (34.7) | 49 (7.1) |  |
| White | 4,785 (34.3) | 803 (28.6) | 517 (75.1) |  |
| Ethnicity (%) |  |  |  | <0.001 |
| Hispanic/Latino | 871 (6.2) | 725 (25.8) | 31 (4.5) |  |
| Non-Hispanic | 8,177 (58.6) | 1,261 (44.9) | 144 (20.9) |  |
| Unknown | 4,898 (35.1) | 821 (29.2) | 513 (74.6) |  |

**Supplementary 3.** Sensitivity analysis with age restriction (minimum required age of 2 years)

**Supplementary Table 1**. Patient and clinical care characteristics of study cohorts aged 0-8 years

|  | **Control** | **ASD** | **Asthma** | **Standardized Mean Difference** |
| --- | --- | --- | --- | --- |
| n | **13,066** | **294** | **1,854** |  |
| Age^a^ | 4.1 (2.1) | 4.9 (1.8) | 5.0 (1.9) | 0.30 |
| Male (%) | 6,131 (46.9) | 230 (78.2) | 1,120 (60.4) | 0.45 |
| Insurance^b^ (%) |  |  |  | 0.22 |
| Private | 10,468 (81.1) | 202 (68.9) | 1,336 (72.4) |  |
| Public | 2,003 (15.5) | 74 (25.3) | 458 (24.8) |  |
| Military | 444 (3.4) | 17 (5.8) | 51 (2.8) |  |
| Race (%) |  |  |  | 0.22 |
| White | 3,882 (29.7) | 67 (22.8) | 474 (25.6) |  |
| Black | 345 (2.6) | 10 (3.4) | 122 (6.6) |  |
| Asian | 2,501 (19.1) | 67 (22.8) | 345 (18.6) |  |
| Other^c^ | 2,066 (15.8) | 59 (20.1) | 404 (21.8) |  |
| Unknown | 4,272 (32.7) | 91 (31.0) | 509 (27.5) |  |
| Ethnicity (%) |  |  |  | 0.15 |
| Hispanic | 1,194 (9.1) | 41 (13.9) | 272 (14.7) |  |
| Not hispanic | 7,440 (56.9) | 160 (54.4) | 1,075 (58.0) |  |
| Unknown | 4,432 (33.9) | 93 (31.6) | 507 (27.3) |  |
| Total Visits^d^ | 11.07 (7.54) | 13.95 (9.64) | 17.31 (11.59) | 0.43 |
| Well-Care Visits^e^ | 5.00 (3.45) | 4.10 (2.93) | 4.17 (2.88) | 0.19 |
| Total Continuity of Care | 0.64 (0.21) | 0.58 (0.21) | 0.57 (0.20) | 0.22 |
| Well-Care Continuity of Care | 0.81 (0.23) | 0.78 (0.23) | 0.80 (0.23) | 0.09 |

^a^Age in years at midpoint in study. Mean and standard deviation is displayed for continuous variables, including age, total visits, well-care visits, same day sick visits, total continuity of care, and well-care continuity of care.

^b^Insurance type in the first visit during the study period.

^c^Includes Other, Native American and Pacific Islander.

^d^Total number of all office visits during the study period.

^e^Number of Well-Care visits during the study period.

**Supplementary Table 2.** Fractional regression models.

| **Total Continuity of Care** | aOR | Lower CI | Upper CI | P value |
| --- | --- | --- | --- | --- |
| Study Cohort (ref=HC) |  |  |  |  |
| ASD | 0.95 | 0.85 | 1.05 | 0.281 |
| Asthma | 0.89 | 0.85 | 0.93 | <0.001 |
| Mean Study Age | 0.99 | 0.99 | 0.99 | <0.001 |
| Total Visits | 0.98 | 0.98 | 0.98 | <0.001 |
| Insurance (ref=Private) |  |  |  |  |
| Public | 0.78 | 0.74 | 0.82 | <0.001 |
| Military | 0.88 | 0.8 | 0.95 | 0.001 |
| Practice (ref=1) |  |  |  |  |
| 2 | 0.73 | 0.68 | 0.77 | <0.001 |
| 3 | 0.6 | 0.54 | 0.66 | <0.001 |
| 4 | 1.92 | 1.82 | 2.02 | <0.001 |
| 5 | 1.55 | 1.49 | 1.61 | <0.001 |
| 6 | 1.05 | 0.99 | 1.11 | 0.099 |
| 7 | 1.75 | 1.7 | 1.8 | <0.001 |
| 8 | 1.82 | 1.7 | 1.93 | <0.001 |
| 9 | 1.64 | 1.56 | 1.72 | <0.001 |
| 10 | 0.52 | 0.41 | 0.62 | <0.001 |
| **Total Continuity of Care** | aOR | Lower CI | Upper CI | P value |
| Study Cohort (ref=HC) |  |  |  |  |
| ASD | 0.98 | 0.82 | 1.13 | 0.751 |
| Asthma | 0.98 | 0.91 | 1.05 | 0.629 |
| Mean Study Age | 1 | 1 | 1 | 0.320 |
| Total Visits | 1.03 | 1.02 | 1.04 | <0.001 |
| Insurance (ref=Private) |  |  |  |  |
| Public | 0.63 | 0.58 | 0.69 | <0.001 |
| Military | 0.91 | 0.77 | 1.06 | 0.218 |
| Practice (ref=1) |  |  |  |  |
| 2 | 0.65 | 0.59 | 0.72 | <0.001 |
| 3 | 0.53 | 0.44 | 0.61 | <0.001 |
| 4 | 4.27 | 3.99 | 4.54 | <0.001 |
| 5 | 1.34 | 1.24 | 1.44 | <0.001 |
| 6 | 1.75 | 1.64 | 1.87 | <0.001 |
| 7 | 2.47 | 2.38 | 2.57 | <0.001 |
| 8 | 1.7 | 1.49 | 1.9 | <0.001 |
| 9 | 1.57 | 1.43 | 1.71 | <0.001 |
| 10 | 0.77 | 0.6 | 0.95 | 0.004 |

ASD=Autism Spectrum Disorder, aOR = Adjusted Odds Ratio, CI = Confidence Interval (95%).
